# Longitudinal Study to Better Understand Neuropathic Back Pain as a Means of Improving Patient Selection for 10 kHz Spinal Cord Stimulation

**DOI:** 10.1101/2024.05.02.24306119

**Authors:** Shreya Mandloi, Christian V Tran, Sara Thalheimer, Samantha Jaffe, Leonard Braitman, Kevin Hines, Ashwini Sharan, Chengyuan Wu

## Abstract

**Objective:** To create a questionnaire that can identify patients who will respond well to high-frequency spinal cord stimulation.

**Materials and Methods:** Thirty-three patients who received permanent implantation of a high-frequency spinal cord stimulator were followed for up to one year. Preoperative patient data in the form of a packet containing pain metrics, the Douleur Neuropathique 4 (DN4) questionnaire, and a questionnaire thought to contain clinically useful questions were collected. Visual analog scale (VAS), Oswestry Disability Index (ODI), and subjective overall percent pain reduction were collected at time points of 6 weeks, 3 months, 6 months, and 12 months.

**Results:** Patients who revealed that they could walk before a long period of time before their pain worsened, whose pain was consistent throughout the day, whose pain wakes them up from sleep and whose pain is worse when leaning side to side had a significant increase in ODI following SCS. Additionally, patients who reported only being able to sit or lie down for less than thirty minutes before having to move experienced significantly decreased ODI.

**Conclusions:** While literature has shown that patients with neuropathic pain respond to SCS, identifying which patients clinically would have an optimal response remains a challenge. Future studies on a greater number of patients are needed to further develop a questionnaire to better identify clinical signs consistent with low back pain that will respond to spinal cord stimulation.

**Statements and Declarations:** *Sources of Financial Support:* Financial support for this project was provided by Nevro

*Conflict of Interests:* Dr. Ashwini Sharan reports personal fees from Neuspera, personal fees from Medtronic, grants and personal fees from Dixi, personal fees and other from Cerebral Therapeutics, outside the submitted work. Dr. Chengyuan Wu reports grants from Nevro, during the conduct of the study; personal fees from Nevro, personal fees from Abbott, personal fees from Medtronic, personal fees from Boston Scientific, personal fees from MicroSystems Engineering, outside the submitted work.

## Introduction

Low back pain (LBP) is one of the most extensive and widespread medical problems, affecting 15-20% of the U.S. population per year.[1] Due to the multifactorial nature of LBP, it is often difficult to treat. It has been proposed that axial lower back pain may consist of three different types of pain: neuropathic (lesion to the somatosensory system itself), nociceptive (lesion to non-neural tissue that activates nociceptors), and nociplastic (gap of patients that don’t have neuropathic or nociceptive pain) pain.[2] However, it is difficult to accurately diagnose and characterize LBP, thus therapeutic responses are often inconsistent. The ability to categorize patients based on what type of axial LBP they have can help clinicians provide more tailored and accurate treatment plans for patients.

The pharmacologic management of neuropathic pain is varied amongst first-line drugs including gabapentin, pregabalin, serotonin-noradrenaline receptor uptakes and tricyclic antidepressants with modest efficacy.[3] Other forms of management for neuropathic pain include physical therapy, psychological therapy, surgery, transcranial magnetic stimulation, and spinal cord stimulation (SCS).[4]Traditional SCS was born out of gate-control theory which predicted improved nociceptive pain as a result of SCS therapy. However, literature has shown that SCS does not adequately impact nociceptive pain and has a stronger impact on neuropathic pain providing relief in greater than 50% of medically refractory neuropathic LBP.[5,6] High frequency (10kHz) SCS is more effective than traditional SCS when used for chronic neuropathic LBP.[7]

Although the specific mechanism of action of high-frequency SCS is not completely known, current theories hypothesize the involvement of both segmental and supraspinal pathways of pain and that SCS may also activate opioid receptors.[8,9] As high frequency SCS has improved and more consistent clinical outcomes in the treatment of neuropathic LBP there is an opportunity to better characterize which patients have neuropathic back and will have optimal response to SCS. There remains a need to identify patients with neuropathic back pain, as no gold standard currently exists and current questionnaires in use have proven insufficient. The Douleur Neuropathique 4 (DN4) questionnaire that is often used was developed for leg pain and is thus not an optimal screening tool for neuropathic back pain.[10-12]As it is difficult to accurately diagnose the type of LBP patients experience, this pilot study aims to assess the utility of screening questions using a physician generated questionnaire to identify patients with neuropathic LBP that respond to high frequency SCS.

## Materials and Methods

### Inclusion/Exclusion Criteria

The study protocol and informed consent were approved by the institutional review board (Study #17C.539) as a prospective before-after clinical trial. Inclusion criteria consisted of patients over 18 years of age who were appropriate candidates for a trial of 10 kHz NEVRO SCS per standard of care. Patients were deemed appropriate candidates if they had failed all conservative measures of pain management including medical management, interventional pain management and physical therapy for at least 6 months. Additionally, patients were deemed appropriate if they had failed prior lumbar spine surgery or as not an appropriate candidate for spine surgery by a board-certified spine surgeon. Patients received either percutaneous or paddle implants at clinical preference of either the patient or the surgeon. Patients younger than 18 years of age or who were unwilling to fill out questionnaires and allow other clinical data to be collected were excluded.

### Questionnaire and Data Collection

A questionnaire was generated based on the implanting physicians’ prior experience as an explicit attempt to differentiate between nociceptive and neuropathic pain. The questionnaire can be visualized in Table I. Participants completed the physician generated questionnaire, Douleur Neuropathique 4 (DN4)[13], Visual Analogue pain scale (VAS)[14], and Oswestry Disability Index (ODI)[15]preoperatively via paper and returned by mail, in-person at appointments, digitally, or by phone call., Flexibility in the way in which patients were allowed to complete the questionnaire was provided in an attempt to increase compliance and retention throughout the study.

**Table I.** Pre-operative questionnaire given to study subjects.

|  |
| --- |
| 1. Is your pain worse with walking? * |
| 2. How long can you walk before your low back pain worsens? ** |
| 3. Did your pain begin all of a sudden (related to a specific event)? * |
| 4. Did your pain gradually get worse as time went on? * |
| 5. How long are you able to stand in one position before having to move? ** |
| 6. How long are you able to sit in one position before having to move? ** |
| 7. How long are you able to lay down in one position before having to move? ** |
| 8. Do you experience pain the entire day? * |
| 9. Is your level of low back pain consistent throughout the day? * |
| 10. Does your pain wake you up from sleep? * |
| 11. After you overexert yourself, does it take more than 1 day to recover? * |
| 12. Is your pain worse immediately when you stand up? * |
| 13. Is your pain improved when leaning forward? * |
| 14. Is your pain worse with bending forward and/or leaning backward? * |
| 15. Is your pain worse with leaning side-to-side? * |
| 16. Does your pain improve after laying down? * |
| 17. How long does it take for the pain to improve after you lay down? ** |
| Response choices: *(yes, no); **(0-5 minutes, 5-20 minutes, 20-30 minutes, >30 minutes) |

(VAS), subjective overall percent pain reduction, and (ODI) scores were collected from patients postoperatively at time points of 6 weeks, 3 months, 6 months, and 1 year. Data was recorded at patient follow-up visits; however, in some patients, data was collected via a phone call due to the COVID-19 pandemic.

### Implantation of Spinal Cord Stimulator

After patients were determined to be candidates for NEVRO SCS therapy and consent was obtained in person in the hospital/clinic, they first underwent a trial with percutaneous electrodes implanted over the T9-T10 interspace for seven to ten days. Patients were considered eligible for permanent implantation if they reported at least a 50% improvement in their pain during their trials, and they underwent permanent implantation one to two weeks after the end of their trials.

### Data Analysis

Longitudinal bivariate mixed effects linear regression models of each ODI, VAS and subjective percent pain reduction as functions of each question were developed separately at each time point (preoperatively, and postoperatively at 1, 3, 6 and 12 months). For each of the bivariate models, variables with p<0.05 were included as potential independent predictors into multivariate mixed models. The questions with p<0.05 were identified as the substantiative questions that were independent predictors of ODI, VAS and subjective % pain reduction.

## Results

### Demographics

A total of 58 patients were enrolled in this study. Of which 25 patients did not fully complete all preoperative baseline measurements and were removed from the overall dataset. Demographic information for the remaining 33 patients (57% compliance) is shown in Table II.

**Table II.** Study Demographics.

| Sex | n | Range | Mean |
| --- | --- | --- | --- |
| M | 11 | - | - |
| F | 22 | - | - |
| Age at Implant | - | 23-84 | 56 |
| <40 | 2 | - | - |
| 40-49 | 6 | - | - |
| 50-59 | 16 | - | - |
| 60-69 | 3 | - | - |
| >69 | 6 | - | - |
| Diagnosis |  |  |  |
| Chronic pain syndrome | 33 | - | - |
| Lumbago with bilateral sciatica | 11 | - | - |
| Lumbago with left-sided sciatica | 4 | - | - |
| Lumbago with right-sided sciatica | 2 | - | - |
| Lumbosacral radiculopathy | 4 | - | - |
| Lumbosacral plexus disorder | 3 | - | - |
| CRPS Type I | 2 | - | - |
| Failed back surgery syndrome | 23 | - | - |
| **Implant Type** |  |  |  |
| Percutaneous | 26 | - | - |
| Paddle | 7 | - | - |

### Questions associated with Changes in ODI

Bivariate analysis was conducted on each question and ODI to find predictors. Through bivariate analysis, question numbers 1-7, 9, 10, and 15 were all significantly associated with changes in ODI and can be seen in Table III. All question numbers with p <0.05 were included in the multivariate analysis except for question 1 which was excluded due to collinearity. Multivariate analysis revealed that patients who could walk for a long period of time before their pain worsened (question 2, p = 0.024), whose pain was consistent throughout the day (question 9, p = 0.049), whose pain wakes them up from sleep (question 10, p = 0.007), and whose pain is worse when leaning side to side (question 15, p = 0.006) had a statistically significantly worsening in ODI following SCS. Additionally, patients that indicated they could stand (question 5, p = 0.006) or sit (question 6, p = 0.002) for a long time in one position before having to move were significantly associated with an improvement in ODI following SCS. All coefficients, confidence intervals and p-values can be seen in Table IV.

**Table III:** Bivariate analysis of Question Number and Changes in ODI.

| Question Number | Coefficient (CI) | p-value |
| --- | --- | --- |
| 1 | 16.95 (1.32, 32.59) | 0.034 |
| 2 | -5.67 (-9.55, -1.78) | 0.004 |
| 3 | 0.500 (0.42, 0.58) | <0.001 |
| 4 | 10.36 (-1.95, 22.66) | 0.099 |
| 5 | -6.68 (-10.11, -3.24) | <0.001 |
| 6 | -7.60 (-10.85, -4.34) | <0.001 |
| 7 | -4.83 (-8.44, -1.21) | 0.009 |
| 8 | -0.88 (-12.04, 10.28) | 0.877 |
| 9 | 8.22 (1.20, 15.24) | 0.022 |
| 10 | 20.04 (6.60, 33.48) | 0.003 |
| 11 | 2.44 (-5.95, 10.82) | 0.057 |
| 12 | 6.22 (-1.83, 14.28) | 0.130 |
| 13 | 6.00 (-1.23, 13.24) | 0.104 |
| 14 | 1.48 (-7.07, 10.02) | 0.735 |
| 15 | 11.08 (3.66, 18.50) | 0.003 |
| 16 | 0.68 (-7.69, 7.82) | 0.986 |
| 17 | 0.34 (-2.89, 3.57) | 0.835 |

**Table IV:** Multivariate Analysis of Question Number and Changes in ODI.

| Question Number | Coefficient (CI) | p-value |
| --- | --- | --- |
| 2 | 11.77 (1.56, 21.98) | 0.024 |
| 3 | -1.27 (-7.82, 5.29) | 0.705 |
| 4 | 11.80 (1.30, 22.30) | 0.028 |
| 5 | -14.73 (-25.14, -4.33) | 0.006 |
| 6 | -5.86 (-9.53, -2.19) | 0.002 |
| 9 | 7.68 (0.02, 15.32) | 0.049 |
| 10 | 22.13 (5.91, 38.34) | 0.007 |
| 15 | 9.39 (2.67, 16.13) | 0.006 |

**Table V:** Bivariate Analysis of Question Number and Changes in % Pain Reduction.

| Question Number | Coefficient (CI) | p-value |
| --- | --- | --- |
| 1 | 9.87 (-10.71, 30.44) | 0.347 |
| 2 | 7.32 (-0.20, 14.84) | 0.056 |
| 3 | -16.94 (-30.65, -3.23) | 0.015 |
| 4 | 7.30 (-20.64, 35.35) | 0.608 |
| 5 | 3.93 (-2.60, 10.47) | 0.238 |
| 6 | 2.95 (-3.57, 9.46) | 0.375 |
| 7 | 2.57 (-4.20, 9.34) | 0.457 |
| 9 | -8.35 (-22.06, 5.36) | 0.233 |
| 10 | -13.81 (-41.96, 14.33) | 0.336 |
| 11 | 32.99 (9.86, 56.12) | 0.005 |
| 12 | -0.20 (-14.80, 14.39) | 0.978 |
| 13 | 5.28 (-8.0, 18.60) | 0.437 |
| 14 | 3.78 (-14.36, 21.92) | 0.683 |
| 15 | -16.86 (-30.79, -2.94) | 0.018 |
| 16 | -11.64 (-24.66, 1.38) | 0.080 |
| 17 | -4.85 (-12.48, 2.77) | 0.212 |

### Questions associated with Changes in Subjective Percent Pain Reduction

Bivariate analysis was conducted on each question and the change in percent pain reduction to find predictors. Question 8 was omitted from bivariate analysis due to collinearity. In bivariate analysis, question numbers 3, 11 and 15 were significantly associated with percent pain reduction and can be seen in Table V. Multivariate analysis revealed that patients who indicated that patients who indicated that their pain is worse when leaning side to side (question 15, p = 0.032) had a significant decrease in subjective percent pain reduction, thus an increase in pain levels following SCS. Coefficients, confidence intervals and p-values for the multivariate analysis can be seen in Table >VI.

**Table VI:** Multivariate Analysis of Question Number and Changes in % Pain Reduction.

| Question Number | Coefficient (CI) | p-value |
| --- | --- | --- |
| 3 | -8.52 (-22.45, 5.41) | 0.231 |
| 11 | 20.87 (-23.64, 65.39) | 0.358 |
| 15 | -15.14 (-28.95, -1.33) | 0.032 |

### Questions associated with Changes in VAS

Bivariate analysis was conducted on each question and VAS to find predictors. Through bivariate analysis, question numbers 5, 9 and 17 were all significantly associated with changes in VAS and can be seen in Table VII. All significant question numbers were included in the multivariate analysis. However, multivariate analysis revealed that no questions were significant independent predictors of changes in VAS and all coefficients, confidence intervals and p-values can be seen in Table VIII.

**Table VII:** Bivariate Analysis of Question Number and Changes in VAS.

| Question Number | Coefficient (CI) | p-value |
| --- | --- | --- |
| 1 | 0.99 (-1.00, 2.99) | 0.330 |
| 2 | -0.52 (-1.06, 0.19) | 0.059 |
| 3 | 0.29 (-0.73, 1.30) | 0.581 |
| 4 | 0.01 (-1.76, 1.78) | 0.991 |
| 5 | -0.50 (-0.98, -0.02) | 0.042 |
| 6 | -0.01 (-0.5, 0.48) | 0.956 |
| 7 | 0.18 (-0.35, 0.71) | 0.509 |
| 8 | -0.05 (-1.51, 1.41) | 0.945 |
| 9 | 1.09 (0.13, 2.04) | 0.026 |
| 10 | 1.69 (-0.16, 3.55) | 0.074 |
| 11 | 0.19 (-0.92, 1.30) | 0.736 |
| 12 | 0.33 (-1.03, 1.10) | 0.952 |
| 13 | -0.68 (-1.65, 0.30) | 0.176 |
| 14 | -0.58 (-1.71, 0.55) | 0.318 |
| 15 | 0.36 (-0.64, 1.37) | 0.482 |
| 16 | -0.05 (-1.12, 1.01) | 0.926 |
| 17 | 0.51 (0.04, 0.98) | 0.035 |

**Table VIII:** Multivariate Analysis of Question Number and Changes in VAS.

| Question Number | Coefficient (CI) | p-value |
| --- | --- | --- |
| 5 | 0.02 (-0.55, 0.59) | 0.939 |
| 9 | 0.67 (-0.41, 1.75) | 0.222 |
| 17 | 0.44 (-0.05, 0.93) | 0.083 |

## Discussion

There is a clinical need to identify patients with neuropathic back pain as no gold standard exists and current questionnaires are often insufficient.[8-12] Separating neuropathic pain from non-neuropathic pain is essential for proper treatment as interventions based on pain mechanisms leads to increased ability to tailor specific therapies.[16]No single sign or physical exam finding is diagnostic of neuropathic pain; thus, it is essential to identify which patients through clinical questionnaires are more likely to have neuropathic pain and respond to SCS.[17]This study aims to lay the groundwork patient clinical characteristics that can classify pain type and predict SCS outcomes.

This study indicates that questions related to a mix of mechanical pain and neuropathic pain were predictive of ODI following SCS. Currently, there is a lack of consensus on the effects of SCS on mechanical pain. Prior studies have demonstrated that SCS can increase pain threshold and decrease mechanical pain sensitivity.[18]Other studies found no difference in mechanical pain following SCS.[19] Our study population demonstrated that patients with mechanical pain – those that responded that their pain worsens when leaning side to side – had a significant worsening in ODI and less pain reduction compared to before SCS. ODI can be used as both a functional measure and to demonstrate pain severity.[20]The functional limitations experienced by patients may be a result of mixed pain. Thus, while SCS has a promising effect on neuropathic pain, ODI can still increase if mechanical pain is a primary issue. Literature demonstrates that pain response is multidimensional and that composite endpoints from multiple pain scales may give more insight into SCS outcome.[21]We additionally found that patients who could sit or stand for a long period of time had improvements in ODI with SCS. These results indicate that patients that can tolerate one position for an increased length of time may be better respondents to SCS than those that must move frequently to relieve their pain. Additionally, patients that could walk for a long time before their pain worsened had significant worsening in ODI with SCS. This highlights that patients that tend to be “better” preoperatively have varying responses to SCS in terms of ODI. Prior literature has demonstrated that the duration of pain does not predict SCS outcome.[22]However, to our knowledge this study emphasizes a clinical gap that preoperative pain level and functionality may influence SCS outcome.

While literature has shown that patients with neuropathic pain respond to SCS, identifying which patients clinically would have an optimal response remains a challenge. Given the significant correlations between specific questions and changes in ODI and subjective pain reduction, this pilot study lays the groundwork for the development of a diagnostic questionnaire to identify appropriate candidates for SCS.

### Limitations

The major limitation of this study is its sample size; as a pilot study, a small number of participants were recruited and so it is possible that different results would be obtained from using a larger dataset. Another limitation is the magnitude of study attrition -- because of the frequency of data collection, low compliance was observed with regards to the questionnaire packets that subjects received at each time point. Additionally, because not all subjects were supervised when filling out the packets, some skipped portions of the packet, resulting in missing data at various time points.

The questionnaire used in this study was also developed solely based on the clinical experience of two authors (CW and ADS). As such, it is unlikely to serve as a comprehensive tool for the identification of subtypes of LBP. At the same time, as this work was developed as a pilot study, the intent of this preliminary questionnaire was to obtain objective feedback on the utility of questions focused on the identification of symptoms consistent with mechanical LBP.

Another limitation is that due to the restrictions of the COVID-19 pandemic, some VAS scores at the latest time point were collected over the phone. As VAS is meant to allow patients to visually represent their perceptions of pain, it is possible that our findings for VAS at that time point are inaccurate.

### Future Studies

Future studies should further assess existing neuropathic pain questionnaires to the one presented in this study with a greater number of participants. Additionally, the results of this study can be used to further develop questions to be presented to participants in future studies, with greater emphasis on questions associated with improved outcomes and the removal of questions that were not predictive. Lastly, further studies are needed to determine the generalizability of these findings to other stimulation paradigms.

## Conclusion

This pilot study attempts to generate clinically relevant questions that can be used to identify patients who experience neuropathic pain in the lower back and are therefore good candidates for high-frequency spinal cord stimulation. Results indicate that questions revealing patients who have a lack of mechanical characteristics are correlated with better outcomes at time points ranging up to one year. Future studies on a greater number of patients are needed to further develop a questionnaire to better identify clinical signs consistent with low back pain that will respond to spinal cord stimulation.

## Data Availability

All data produced in the present work are contained in the manuscript

